# Real-world feasibility of ERS asthma diagnosis guidelines for school-aged children

**DOI:** 10.1101/2025.09.12.25335571

**Authors:** Mari Sasaki, Myrofora Goutaki, Sarah Glick, Sylvain Blanchon, Karin Hoyler, Philipp Latzin, Alexander Moeller, Nicolas Regamey, Claudia E. Kuehni, SPAC Study Team

**Author notes:** **Corresponding author:** Prof. Claudia E. Kuehni, Institute of Social and Preventive Medicine, University of Bern, Mittelstrasse 43, 3012, Bern, Switzerland.

## Abstract

**Background:** Clinical practice guidelines for asthma diagnosis are rarely evaluated in real-life practice. Within the Swiss Paediatric Airway Cohort (SPAC), we initiated the SPAC-asthma project to develop a standardised diagnostic approach for school-aged asthma, based on the algorithm recommended by the European Respiratory Society (ERS) guideline. Here, we report the development and feasibility of this approach after implementation across multiple paediatric pulmonology clinics.

**Method:** We used a modified Delphi process with paediatric pulmonologists from participating clinics to tailor the ERS algorithm for feasible implementation in children aged 5-17 years with suspected asthma. Key adaptations included selection of initial tests, criteria for further testing, test cutoffs, the role of medication trial and follow-up procedures. One year after implementation, we evaluated adherence to the adapted approach at four clinics and explored the reasons for any deviations.

**Results:** The final SPAC-asthma approach included spirometry, fractional exhaled nitric oxide and allergy testing as initial tests, followed by either bronchodilator reversibility testing, bronchial challenge test or medication trial. Overall adherence after one year was 77% (182/236 patients). Deviations were due to practice-related (e.g., different criteria for bronchial obstruction), patient-related (e.g., inability to perform spirometry), and logistical reasons (e.g., scheduling difficulties).

**Conclusion:** The diagnostic approach was well implemented, but the observed deviations highlighted the need for flexibility when applying guidelines in real-world settings. As a next step, we will assess whether implementing the ERS asthma guidelines in school-aged children improves diagnostic accuracy.

**Take home message:** We tested a standardised ERS guideline-based approach to diagnose school-age asthma across Swiss paediatric pulmonology clinics. After expert adaptation and a year, adherence was good and we identified areas to improve guideline implementation.

## Introduction

The importance of implementing clinical guidelines is increasingly recognised (1–3). However, the feasibility of these guidelines in real-world settings is often not adequately assessed or addressed during development (4–6). This also applies to asthma, where current guidelines recommend standardised diagnostic approaches that combines several tests to reduce misdiagnosis (7–9). Yet, there is limited real-world evidence on the feasibility and accuracy of these approaches in school-age children.

In 2020, we retrospectively analysed data from 514 children suspected of asthma in the Swiss Paediatric Airway Cohort (SPAC) to evaluate asthma diagnostic algorithms proposed by National Institute for Health and Care Excellence (NICE) and the Global Initiative for Asthma (GINA) guideline (7, 8, 10). When comparing the diagnoses given by paediatric pulmonologists with those suggested by the guideline algorithms using the tests performed, both showed suboptimal accuracy. In addition, the NICE algorithm was applicable in only 17% of cases due to its reliance on peak expiratory flow rate (PEFR) variability, which is not commonly used in Switzerland. These results raised concerns about the performance of these guidelines in the Swiss setting and highlighted the limitations of retrospective analyses in multi-centre studies, in which not all patients undergo the same diagnostic tests.

In 2021, the European Respiratory Society (ERS) clinical practice guidelines for the diagnosis of asthma in children aged 5-16 years were published (11). This guideline was based on an extensive systematic review of evidence on the individual tests, and included a diagnostic algorithm developed through a modified Delphi process, informed by available evidence, expert consensus and practical considerations. The algorithm (further referred to as “ERS algorithm”) combines spirometry, bronchodilator reversibility (BDR) testing, fractional exhaled nitric oxide (FeNO) measurement and bronchial challenge tests, PEFR or medication trials when needed. In 2022, Swiss recommendations on asthma diagnosis were published proposing an adapted algorithm adjusting the ERS algorithm for use in Switzerland by removing PEFR and adjusting the BDR positivity threshold to a 10% increase in percent predicted FEV_1_, reflecting updated technical standards (12, 13).

To assess the proposed diagnostic approach in the ERS guideline, we initiated the SPAC-asthma project in 2022, supported by the Swiss National Science Foundation. This project comprised three phases: (1) development of a standardised diagnostic approach based on the ERS guideline, (2) implementation and evaluation of its feasibility across paediatric pulmonology clinics, and (3) assessment of its diagnostic accuracy. This paper presents results from phases 1 and 2: the development of the approach and its feasibility after one year, based on adherence and reasons for deviations.

## Material and methods

### Swiss Paediatric Airway Cohort (SPAC)

The SPAC-asthma project is embedded within SPAC, an ongoing Swiss multi-centre clinical observational study. SPAC includes 0–17-year-old children referred to paediatric pulmonologists for their respiratory problems, such as cough, wheeze and exercise induced respiratory symptoms. Details of the cohort have been published previously (10, 14, 15). In SPAC, we collect clinical data such as diagnoses and test results from medical records, and symptoms and demographic information through questionnaires at baseline and annually thereafter. We obtained ethical approval from the Bernese ethics committee (KEB 2016–02176) and all participating parents and adolescents aged 14 years or older gave written informed consent.

Children with suspected asthma are usually referred by their primary care paediatricians in Switzerland, as diagnostic testing is often unavailable in primary care. At the 10 participating SPAC centres, the initial diagnostic work-up for school-aged children with suspected asthma typically includes clinical evaluation, FeNO measurement, spirometry or body plethysmography including spirometry, BDR when indicated, and allergy testing for phenotyping, not necessarily for diagnosis. Bronchial challenge tests are conducted when asthma diagnosis remains inconclusive, either during the same or at a later appointment. Appointment scheduling is often constrained by extended waiting times for specialist consultations.

### SPAC-asthma project study population

The SPAC-asthma project aimed to standardise the asthma diagnostic approach across clinics, with minor adaptations to the ERS algorithm based on local test availability, routine practices, and appointment scheduling. We invited paediatric pulmonology clinics already participating in SPAC to take part in both the development and implementation of the approach in their routine clinical practice. Five clinics joined the project: three university hospitals (Bern, Zurich and Lausanne), one cantonal hospital (Lucerne), and one private practice (Horgen). Participating centres applied the standardised approach to all eligible children, in contrast to previous SPAC procedures, where diagnostic testing varied between clinics.

We included children in the SPAC-asthma project if they were aged 5–17 years, enrolled in SPAC through one of the participating clinics, and newly referred with suspected asthma or asthma-like symptoms after the implementation of the standardised diagnostic approach in January 2024.

#### Phase 1: Delphi process for developing a standardised approach for asthma diagnosis

To develop the standardised diagnostic approach, we used a modified Delphi process with an expert panel of six experienced paediatric pulmonologists: the heads of the five participating SPAC-asthma clinics and the project’s principal investigator. We defined a consensus as ≥80% agreement within the panel. Additional SPAC collaborators and the study team contributed to discussions but did not vote.

The Delphi process included three steps (Figure 1): (1) preliminary group meetings, (2) individual semi-structured interviews and (3) consensus-building group meetings. Discussions focused on: tests routinely performed at the initial visit, criteria for second-step tests (e.g., BDR and bronchial challenge tests), test procedures and cutoffs, the role of medication trial, and follow-up visit procedures. Between December 2022 and June 2023, we held three preliminary meetings to introduce the project and to identify differences in clinical practice across centres and with the ERS guideline. In July and August 2023, we conducted individual semi-structured interviews with each centre head to further understand diagnostic practices and assess the feasibility of the standardised approach. Findings from the interviews guided three follow-up group meetings, during which the approach was refined, and consensus was reached. In November 2023, the expert panel achieved 100% agreement on the final diagnostic approach.

**Figure 1.**
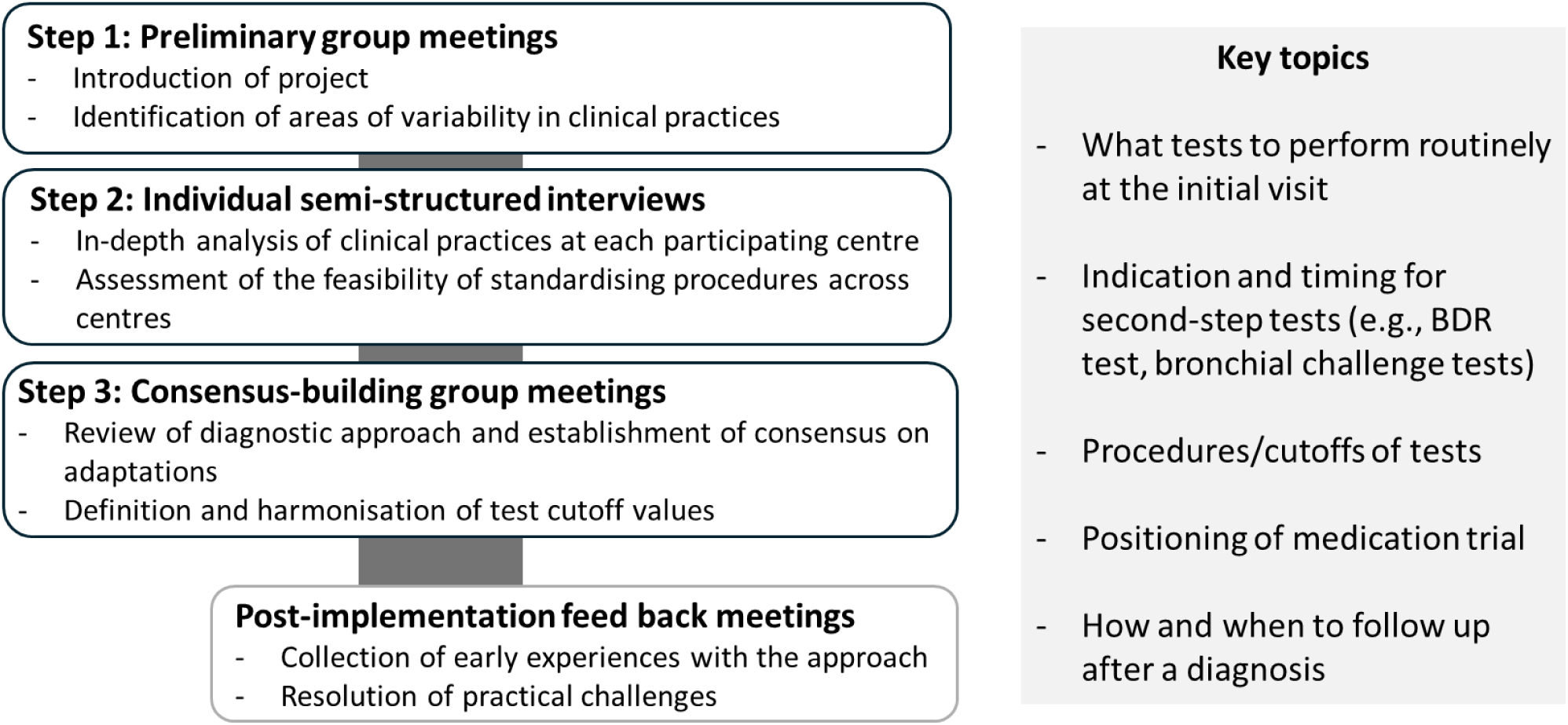
SPAC-asthma project phase 1: Delphi process to develop and implement a standardised diagnostic approach for asthma in school-age children Abbreviations: BDR= bronchodilator reversibility

#### Phase 2: Implementation and feasibility analysis after one year

The standardised diagnostic approach was implemented in the participating clinics in January 2024. Shortly after implementation, the study team held group discussions with the heads of the SPAC-asthma clinics to assess early experiences with the approach, address practical challenges, and refine details such as test procedures and visit intervals.

To assess feasibility after one year, focusing on the diagnostic work-up visits, we analysed data from children who participated in the SPAC-asthma project during 2024 at four of the participating clinics (Bern, Horgen, Lucerne and Zurich). The fifth clinic was excluded from the analysis due to the limited number of enrolled patients, resulting from the clinic’s relocation during the study period. For each child, we reviewed whether diagnostic tests were conducted and interpreted following the standardised approach implemented at that centre. We measured adherence to the approach as the proportion of children who followed the tests described in the approach. For cases with deviations, we identified and categorised the underlying reasons to better understand barriers to consistent implementation.

## Results

### Phase 1: Development of the standardised diagnostic approach

#### 1. Final diagnostic pathways

The Delphi process resulted in a standardised diagnostic approach with two pathways, slightly differing in when BDR was performed and how it was interpreted (Figure 2).

- Both pathways: Initial visit includes spirometry (body plethysmography as optional), FeNO, and allergy testing.
- Pathway 1: All children receive BDR, with a positive result defined as ≥ 12% increase in FEV_1_. If BDR is negative and clinical suspicion remains, a bronchial challenge test (methacholine challenge test or exercise challenge test) is performed at the second visit, or a medication trial follows.
- Pathway 2: BDR is performed if spirometry is obstructive, with a positive result defined as ≥ 10% increase in percent predicted FEV_1_. Children with normal spirometry receive a challenge test (or initiation of medication trial) during the same visit. Those with a negative BDR and ongoing suspicion receive a challenge test at a separate visit or a medication trial.

**Figure 2.**
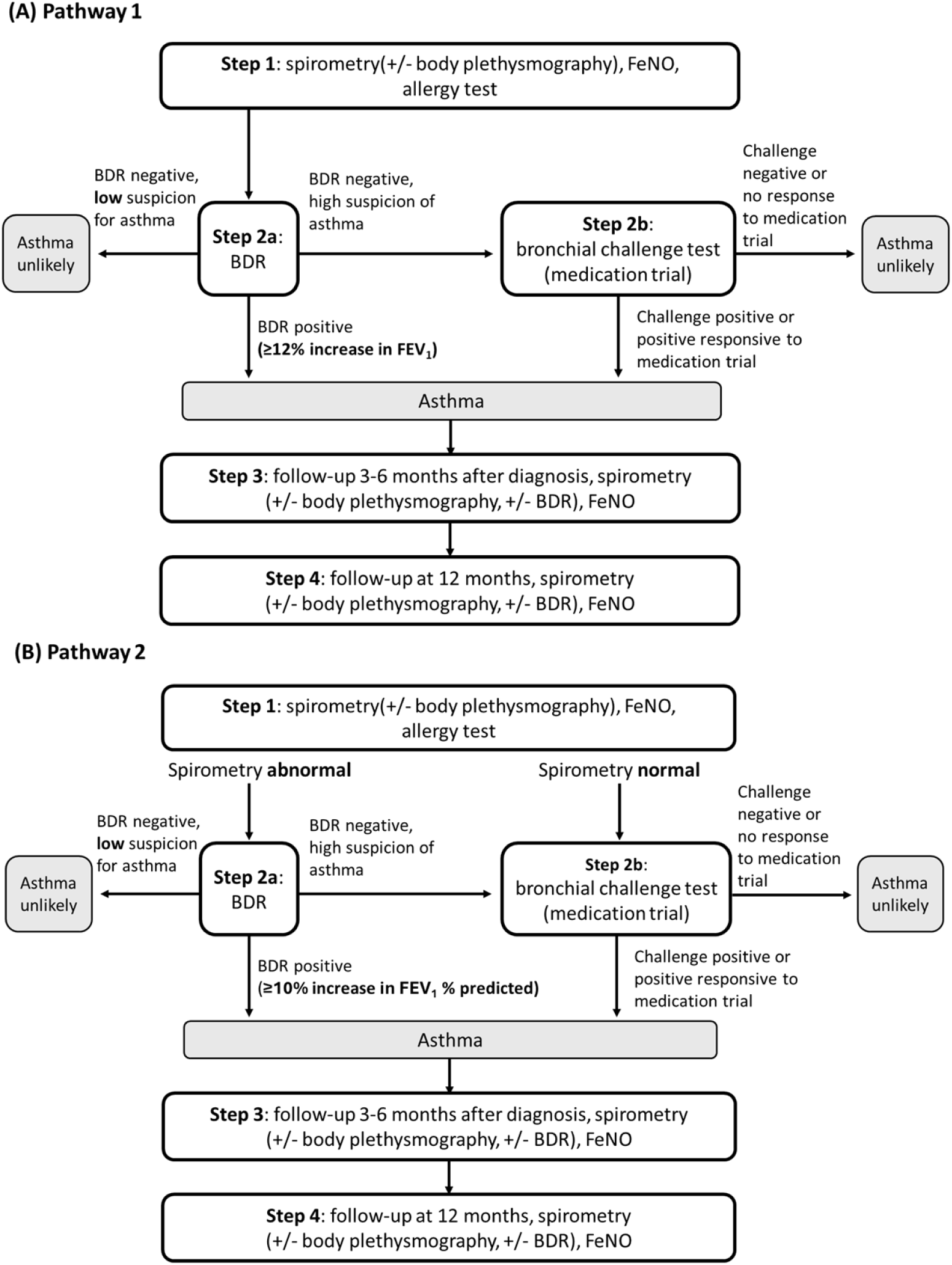
SPAC-asthma project: the two pathways included in the standardised diagnostic approach for asthma among school-age children in Switzerland. * Bronchial challenge test includes methacholine challenge test and exercise challenge test. Medication trial is recommended when challenge test is not feasible. It is defined as a re-assessment at 3-6 months after treatment using inhaled corticosteroid (ICS) alone or inhaled ICS and long-acting β2-agonist (LABA) in combination (ICS-LABA). A positive response to medication trial is >7% increase in prebronchodilator FEV1 plus clinical improvement. The re-assessment will include BDR when possible. Abbreviations: BDR = bronchodilator reversibility; FeNO = fractional exhaled nitric oxide; FEV₁ = forced expiratory volume in 1 second

#### 2. Key discussion points and adaptation decisions (Table 1)

##### First-step routine tests

Spirometry, FeNO, and allergy testing were agreed upon as first-step routine assessments. FeNO was to be measured before spirometry in all cases, regardless of spirometry results.

**Table 1.** Main discussion points and adaptation decisions in the SPAC-asthma project for the - implementation of the ERS diagnostic algorithm.

|  | **ERS guideline recommendation (11)** | **Clinical practice and discussions** | **Final adaptation decisions** |
| --- | --- | --- | --- |
| **First-step routine tests** |  |  |  |
| **Which tests should be done routinely in which order at the initial visit?** | Spirometry, BDR and FeNO recommended as first-line tests, FeNO positioned after spirometry. | Allergy testing routinely done for phenotyping; body plethysmography often used. | Spirometry, FeNO (before spirometry), and allergy testing included as standard; body plethysmography optional. |
| **Second-step tests** |  |  |  |
| **Who should undergo BDR?** | Performed only if obstruction seen on spirometry. | Some centres performed BDR routinely, regardless of spirometry results; some only when obstruction was seen due to logistics or diagnostic preference. | Two diagnostic pathways created: one with universal BDR, one conditional on obstruction. |
| **Who should undergo a bronchial challenge test?** | Recommended if spirometry is normal and FeNO $\geq 25$ ppb. | Conducted in unclear cases after spirometry (with or without BDR) regardless of the FeNO value. | Indication broadened to include all cases with unclear diagnosis after spirometry or BDR. |
| **Test procedures, interpretations and cutoffs** |  |  |  |
| **What is an abnormal spirometry (i.e., airway obstruction present)?** | FEV<sub>1</sub>/FVC < LLN or < 80% predicted or FEV<sub>1</sub> < LLN or < 80% predicted. | Flow-volume curve shape also considered for obstruction. | Concave flow-volume curve added as a criterion for bronchial obstruction |
| **What is the cutoff a positive BDR?** | $\geq 12\%$ increase in FEV<sub>1</sub>. | Some clinics preferred $\geq 10\%$ increase in % predicted FEV<sub>1</sub> based on the latest technical standard (13). | Both cutoffs used in different pathways to accommodate preferences. |
| **How should allergy tests be conducted?** | Not specified. | Methods (e.g., skin prick test, specific IgE) and test validity period varied. | Clinics retained discretion; minimum allergen panel defined; validity set at 2 years (up to 3 in exceptions) |
| **How should methacholine challenge test be conducted and interpreted?** | Positive if PC20 $\leq 8$ mg/ml. | Protocols varied; full harmonisation not feasible. | Agreement to continue discussion toward standardisation. |
| **Medication trial** |  |  |  |
| **Who should receive a trial of medication?** | Not considered a diagnostic tool; recommended if BDR negative and FeNO ≥ 25 ppb. | Commonly used when challenge test not feasible (due to age, technique, scheduling, or parent refusal). | Accepted as alternative when challenge test not feasible. |
| **How should response to medication trial be evaluated?** | Review after 1–2 months; assess symptoms and spirometry (≥7% FEV<sub>1</sub> improvement). | 1–2 months often not feasible due to scheduling. | Review interval extended to 3–6 months with symptom and spirometry assessment; BDR when possible. |
| **Follow up visits** |  |  |  |
| **When should children be followed up after an asthma diagnosis?** | Not specified. | 3–6 months and/or 12 months, depending on clinic/case. | Standardised at 3–6 months and 12 months post-diagnosis. |
| **Which tests should be done at the follow up visit?** | Not specified. | Spirometry (± BDR) and FeNO commonly performed. | Spirometry and FeNO set as routine; body plethysmography and BDR optional. |

Abbreviations: BDR = bronchodilator reversibility; FeNO = fractional exhaled nitric oxide; ICS = inhaled corticosteroid; LLN = lower limit of normal; PC20 = provocative concentration causing 20% fall in FEV<sub>1</sub>.

##### Second-step tests

One key area of disagreement was the indication for BDR. Some clinics routinely performed BDR in almost all children, based on the potential diagnostic value even without airway obstruction (16). Others preferred limiting BDR to children with obstructive spirometry, in line with the ERS algorithm, due to clinical preference and logistical difficulties with scheduling another appointment for bronchial challenge tests if BDR was negative. As a solution, we developed two pathways: Pathway 1, with BDR testing for all children; and Pathway 2, with BDR testing only for those with abnormal spirometry. Different to the ERS algorithm, the indication for bronchial challenge test was expanded to be independent of FeNO values. This decision reflected variation in expert opinion regarding the role of FeNO, whether primarily as a phenotyping or diagnostic tool, and the practical capacity of centres to conduct challenge tests.

##### Test procedures, interpretations and cutoffs

Final decisions are summarised in Table 2 with details on test procedures provided in the supplementary material. In addition to the ERS criteria using FEV_1_ and FEV_1_/FVC in the interpretation of spirometry, a concave flow-volume curve was accepted to indicate bronchial obstruction (17–19). In response to the differing opinions on the BDR cutoff, we used ≥12% increase in FEV_1_ in Pathway 1 and ≥10% increase in percent predicted FEV_1_ in Pathway 2 (11, 13). Allergy test results from any method within the past 2 (exceptionally 3) years were accepted, with house dust mite, birch, grass, cat, and dog dander defined as the minimum allergen panel. Harmonisation of the methacholine challenge procedure remained incomplete due to variations in clinic protocols and equipment, and discussions are ongoing.

**Table 2.**
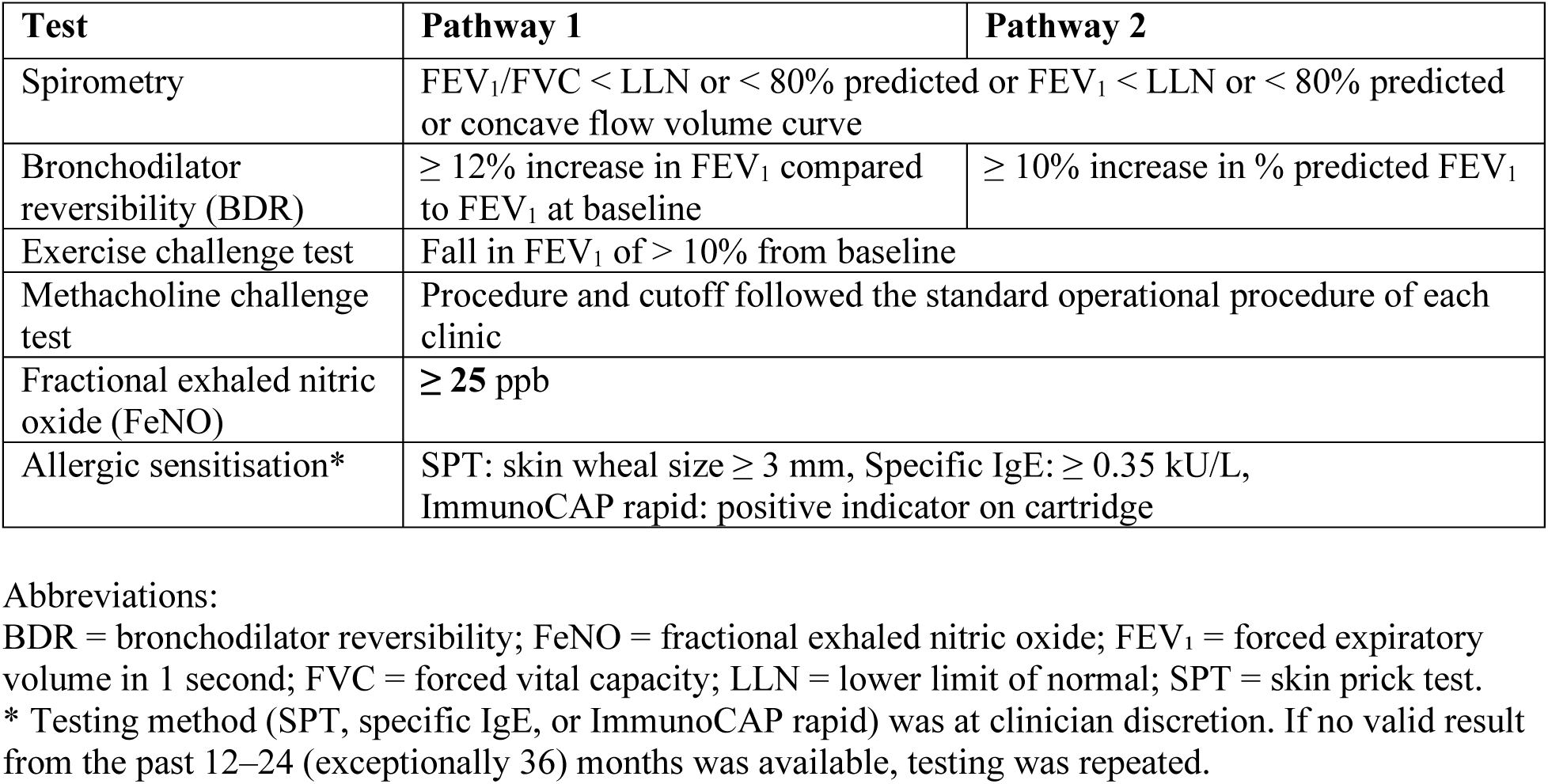
Definition of positive diagnostic test results in the SPAC-asthma approach for children aged 5–17 years.

##### Medication trial

A medication trial with re-evaluation was included as an alternative step when a challenge test was not feasible. Re-evaluation required not only symptom frequency but also spirometry, and a BDR when feasible. A >7% increase in pre-bronchodilator FEV_1_ was considered as a significant improvement in the ERS guideline (11). The interval of assessment was extended to 3–6 months to accommodate scheduling constraints.

##### Follow up visits

We standardised the follow-up visits at 3–6 months and 12 months after diagnosis, with spirometry and FeNO as routine tests. Although a 3-month follow-up was clinically preferred, this was not feasible in several clinics due to scheduling constraints.

### Phase 2: Feasibility analysis after one year of implementation

Of 252 children aged 5-17 years enrolled in SPAC through the four participating clinics in 2024 with clinical data available, 16 were enrolled at follow-up or had previously diagnosed asthma. Thus, 236 met the inclusion criteria for the SPAC-asthma project. The median age was 9 years (interquartile range 7 to 11) and 53% were male.

#### 1. Adherence to the diagnostic approach

Among the 236 children, 97 were evaluated at two clinics implementing Pathway 1 and 139 at two clinics using Pathway 2. Figure 3 shows the number of children at each step. All received initial routine tests, except for three children who missed allergy tests. The overall adherence was 81% (79/97) for Pathway 1 and 74% (103/139) for Pathway 2.

**Figure 3.**
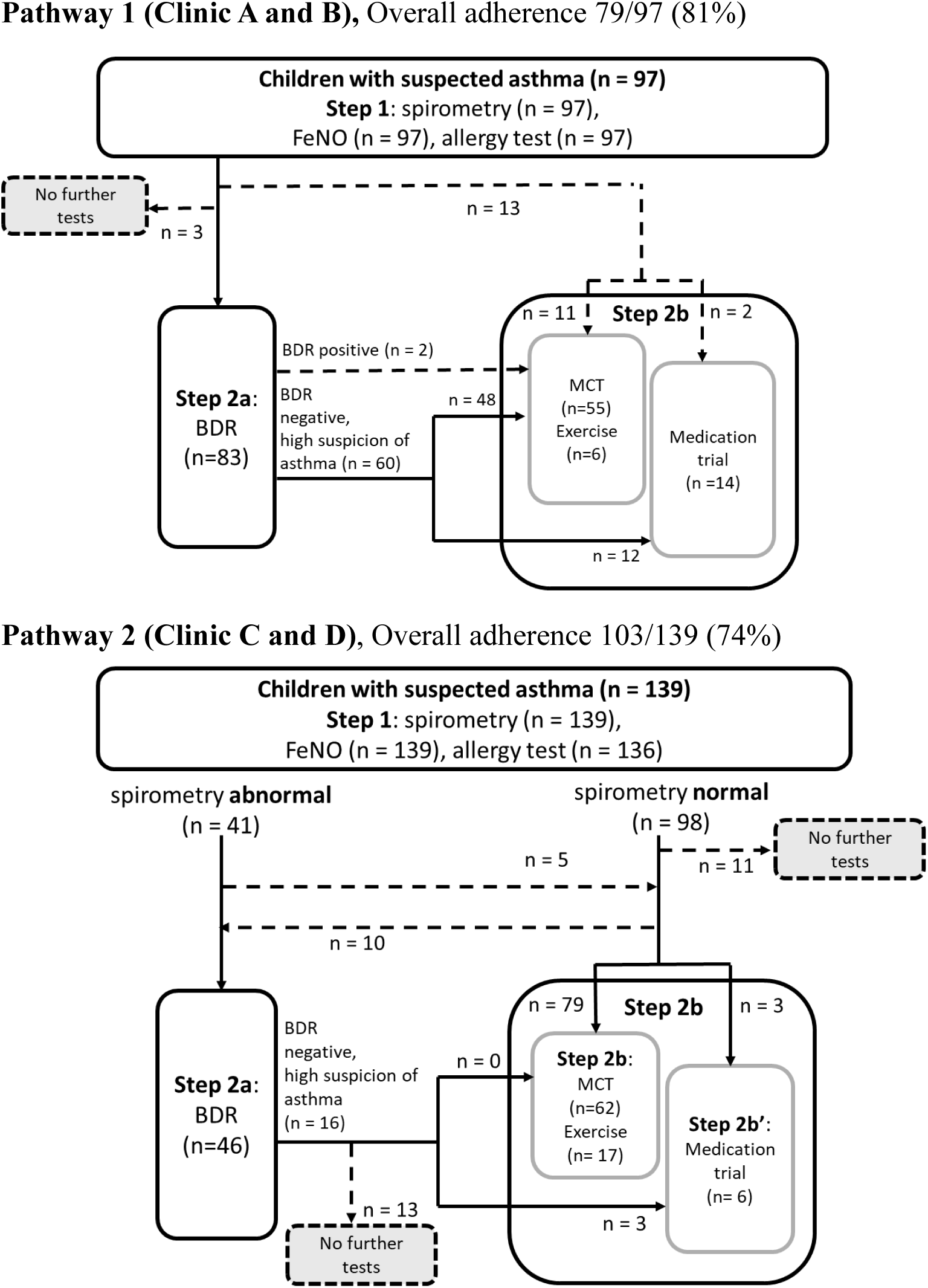
Adherence to each diagnostic pathway implemented across the four clinics participating in the SPAC-asthma project Dotted lines describe deviations from the pathways. Abbreviations: BDR = bronchodilator reversibility; FeNO = fractional exhaled nitric oxide, MCT = methacholine challenge test; Exercise = exercise challenge test

Deviations in Pathway 1:

- BDR was skipped in favour of a challenge test or medication trial (n=13).
- No second-step test performed (n=3).
- Challenge test performed despite a positive BDR (n=2).

Deviations in Pathway 2:

- No further testing after negative BDR despite ongoing suspicion (n=13).
- No second-step test performed (n=11).
- BDR performed despite normal spirometry (n=10).
- Challenge test done instead of BDR after abnormal spirometry (n=5).

#### 2. Reasons for deviations

We identified the reasons in 54 of the 57 deviations based on the medical records (Table 3 and Supplementary table 1) and grouped them into the following three categories.

**Table 3.**
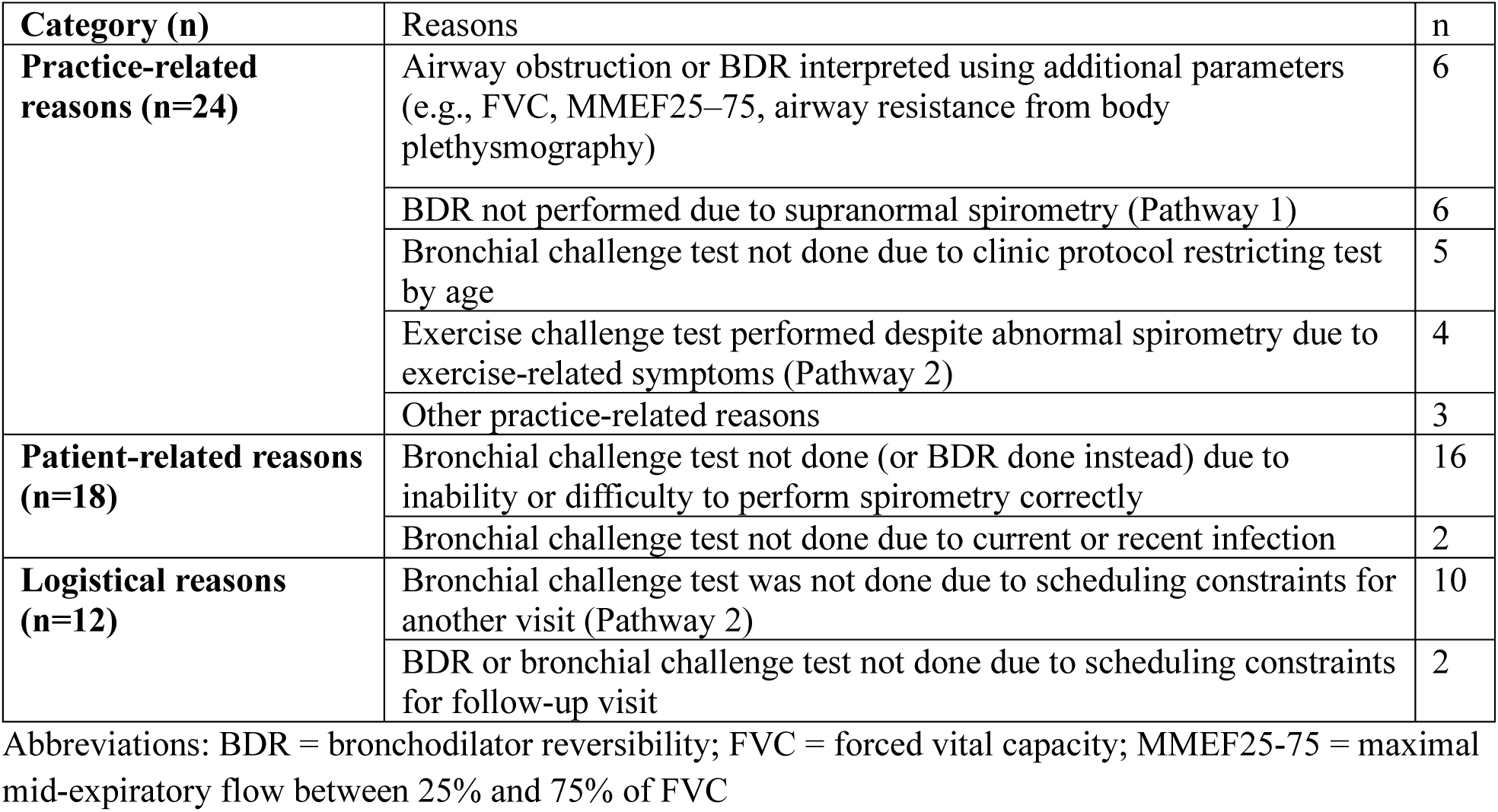
Reasons for deviations from the standardised diagnostic approach in the SPAC-asthma project.

| Category (n) | Reasons | n |
| --- | --- | --- |
| **Practice-related reasons (n=24)** | Airway obstruction or BDR interpreted using additional parameters (e.g., FVC, MMEF25–75, airway resistance from body plethysmography) | 6 |
|  | BDR not performed due to supranormal spirometry (Pathway 1) | 6 |
|  | Bronchial challenge test not done due to clinic protocol restricting test by age | 5 |
|  | Exercise challenge test performed despite abnormal spirometry due to exercise-related symptoms (Pathway 2) | 4 |
|  | Other practice-related reasons | 3 |
| **Patient-related reasons (n=18)** | Bronchial challenge test not done (or BDR done instead) due to inability or difficulty to perform spirometry correctly | 16 |
|  | Bronchial challenge test not done due to current or recent infection | 2 |
| **Logistical reasons (n=12)** | Bronchial challenge test was not done due to scheduling constraints for another visit (Pathway 2) | 10 |
|  | BDR or bronchial challenge test not done due to scheduling constraints for follow-up visit | 2 |

Abbreviations: BDR = bronchodilator reversibility; FVC = forced vital capacity; MMEF25-75 = maximal mid-expiratory flow between 25% and 75% of FVC

Practice-related reasons: These included clinic- or physician-specific decisions, such as using additional parameters for the interpretation of bronchial obstruction and BDR (e.g., FVC, MMEF_25–75_) or setting age or symptom-based criteria for the indication of tests.

Patient-related reasons: These were mainly the difficulty or inability to perform adequate spirometry in young children leading to deviations, such as choosing BDR over bronchial challenge tests due to fewer required manoeuvres.

Logistical reasons: Scheduling constraints, especially difficulty for bronchial challenge test after a negative BDR in Pathway 2, led to deviations from the pathway.

## Discussion

We prospectively implemented a standardised diagnostic approach for asthma in school-age children, based on the ERS guideline, across multiple Swiss paediatric pulmonology clinics in secondary and tertiary care. We adapted the ERS algorithm through a modified Delphi process over a period of more than 12 months to reflect real-world clinical practices and improve feasibility. The resulting approach included two pathways. After one year, we observed good overall adherence and identified several practice-related, patient-related and logistical factors that offer important insights for future guideline development and implementation.

Our results reflect the setting of the SPAC-asthma project, which was conducted in paediatric pulmonologist clinics, primarily within hospitals. Compared to primary care or non-specialist settings, these clinics have greater access to diagnostic resources and clinical expertise, on the other hand, they face challenges such as limited appointment availability. This setting influenced several adaptations to the ERS algorithm, including broader use of BDR (in Pathway 1) and bronchial challenge tests, use of flow-volume curves to assess obstruction, and extended follow-up intervals after medication trials. Many of the deviations from the pathways also reflect this specific clinical setting. Practice-related deviations occurred when specialists applied their clinical expertise, such as using additional lung function parameters or considering symptoms in selecting tests. Logistical reasons, for example, limited scheduling capacity for second-step testing, also led to deviations.

Patient-related deviations from the SPAC-asthma pathway were mostly due to the inability to perform a reliable spirometry test, often in younger children. Although all participating clinics had trained staff, the procedure remains more challenging in 5-7 year old children (20). This points to both the difficulty of confirming asthma in this age group using spirometry and spirometry-dependent tests like BDR and bronchial challenge tests. While such limitations cannot be completely avoided, repeated assessments over time may help improve diagnostic certainty (21, 22). Repeated assessments may also be relevant for children with persistent clinical suspicion after negative test results, for example when prior ICS treatment may have influenced the test results (23). Such situations are not fully addressed in current algorithms and may warrant consideration in future guideline adaptations.

Our findings highlight that guidelines are designed for broad contexts and typical patient presentations, yet implementation requires adaptation to the setting and to variable patient presentations (2, 24, 25). This need should be considered in guideline development and in clinical use. We summarised the main takeaway messages for guideline development and implementation in Text box 1. More such studies are warranted to explore feasibility in diverse healthcare settings and to identify context-specific adaptations or challenges. In addition, some recommendations within guidelines may rest on limited evidence, such as for several diagnostic tests in the ERS guideline (11). Implementation studies can therefore not only test feasibility but also help strengthen the evidence base for such recommendations.

This study is unique in adapting the guideline as part of the implementation and evaluating real-world feasibility. Guideline implementation is increasingly recognised as important across health care (3, 4, 24–27). However, most published studies focus on strategies to improve uptake or barriers to use rather than feasibility in practice (1, 2, 28, 29). In paediatric asthma, several studies have tested interventions in hospital settings aimed at improving adherence to guideline-recommended treatment of acute exacerbations or examined adherence to guidelines using surveys (30–36). By contrast, we explicitly tested the real-world feasibility of applying the diagnostic guideline and identified challenges and adaptations required in Swiss paediatric pulmonology clinics. A further strength was the active involvement of clinic heads in adapting the algorithm and developing the diagnostic approach, which helped address potential reasons for deviation that might otherwise have been overlooked.

Several limitations should be considered. First, the specific adaptations and challenges we found may not be directly transferable to other contexts. Although the approach was standardised, it is mainly applicable to settings with access to FeNO, spirometry, and bronchial challenge tests. Diagnostic processes may differ considerably in primary care, where such tests are often less readily available. Second, although the study implemented diagnostic pathways prospectively, deviations were assessed retrospectively, which may have introduced some misclassification or limited our ability to fully capture the underlying reasons for deviations. Finally, we conducted the project in five clinics and involved a relatively small number of children, which may have reduced the level of detail we could explore in the analysis.

In summary, our findings show that with active clinician engagement, implementing a guideline-based diagnostic approach for school-age asthma is feasible across multiple centres. The necessary adaptations and observed deviations highlight the need for flexibility in future guideline use and for continued evaluation in clinical practice. As the next step in this project, we will evaluate the diagnostic accuracy of the adapted pathways. This will provide valuable insights into the effectiveness of guideline-based algorithms in real-world clinical settings.

#### Text Box 1. Suggestions for guideline development and implementation

##### For guideline development

➢ Incorporate considerations for real-world implementation, including variability in clinical settings and resource limitations.
➢ Promote and support prospective feasibility studies in diverse healthcare contexts.

##### For guideline implementation in practice

➢ Account for setting-specific factors such as diagnostic test availability, interpretation differences, and logistical constraints.
➢ Identify and address patient groups requiring tailored approaches (e.g., younger children unable to perform reliable spirometry).
➢ Work with colleagues and the clinical team to identify necessary adaptations, ensure feasibility and a common understanding of how to apply the guideline.
➢ Conduct feasibility studies within your setting and communicate the findings, thereby contributing to refinement and improvement of future guidelines.

## Acknowledgements

The authors would like to thank the children and families who took part in the SPAC study and the members of the SPAC Study Team. Members of the current and past SPAC Study Team are: T. Schürmann and C. Bieli (Cantonal Hospital Aarau, Aarau, Switzerland); A. Jochmann, D. Trachsel and J. Usemann (University Children’s Hospital Basel, Basel, Switzerland); P. Latzin, C. Casaulta, M. Bullo, I. Korten, E. Kieninger, B. Frauchiger, B. Vomsattel, S. Yammine and C.C.M de Jong (Inselspital, Bern University Hospital, University of Bern, Bern, Switzerland); P. Iseli (Cantonal Hospital Graubünden, Chur, Switzerland); K. Hoyler (private paediatric pulmonologist, Horgen, Switzerland); S. Blanchon, S. Guerin, I. Rochat and C. Fernandez-Elviro (Lausanne University Hospital, University of Lausanne, Lausanne); N. Regamey, M. Lurà, M. Hitzler, K. Hrup, E. Sidler, P. Heer and C. Bernold (Children’s Hospital of Central Switzerland, Lucerne, Switzerland); J. Barben (Children’s Hospital St. Gallen, St. Gallen, Switzerland); O. Sutter and L. Krüger (private paediatric practice, Worb, Bern, Switzerland); A. Moeller, E. Seidl, G. Signorelli, YT. Lam, G. Buggle, S. Beck and L. Benz (University Children’s Hospital Zurich, Zurich, Switzerland); and C.E. Kuehni, M. Goutaki, C. Ardura-Garcia, D. Berger, S. Glick, B. Guerra, T. Krasnova, R. Makhoul, M.C. Mallet, E. Pedersen, F. Romero, M. Sasaki and V. Weihrauch (Institute of Social and Preventive Medicine, University of Bern, Bern, Switzerland).

## Author contributions

CK, SG, and MG conceptualised the study. SB, KH, PL, AM and NR recruited patients and took part in the investigation. MS conducted the analysis. CK and MG supervised the research. MS drafted the manuscript, and all authors contributed to its revision. All authors approved the final version of the manuscript.

## Data availability statement

SPAC study data is not deposited at an open access repository, as participants were not asked to give consent to have their data deposited publicly. Requests for partial datasets for specific analyses including individual patient data and a data dictionary defining each included field can be addressed to Prof Kuehni upon reasonable request.

