## Supplementary material for "Real-world feasibility of ERS asthma diagnosis guidelines for school-aged children"

**Details on test procedures for the four SPAC-asthma project clinics**

*Spirometry and bronchodilator reversibility*

Spirometry was performed with a MasterScreen Pneumo spirometer (Vyaire Medical, Chicago, IL, USA) using Sentrysuite software or SMART PFT BODY (Lemon Medical Gmbh, Hammelburg, Germany) in accordance with the ERS/ATS technical standards (1). Experienced lung function technicians or nurses performed quality control during the process and record the best measurement of the three trials. Reference values were based on the Global Lung Function Initiative 2012 equations (2). Bronchodilator reversibility was assessed by the change in lung function, 10-15 minutes after administration of salbutamol 400 μg by pressurised metered-dose inhaler via spacer.

*Fractional exhaled nitric oxide (FeNO)*

FeNO by single-breath online method was measured using ANALYZER CLD 88 sp (Eco Medics AG, Duernten, Switzerland) in accordance with ATS/ERS recommendations (3). Measurement was done before spirometry in all centres and the mean value from two measurements was assessed.

*Bronchial challenge tests*

Exercise challenge tests were conducted using a cycle ergometer or motorized treadmill. Spirometry was repeated after completion of sufficient exercise and results were described as the maximum fall in FEV1 compared to the baseline. Methacholine challenge tests were carried out using the Vyntus Aerosol Provocation System (Vyaire Medical, Chicago, IL, USA) or SMART PFT NEBULIZER (Lemon Medical Gmbh, Hammelburg, Germany), and the dose protocol was 50, 50, 200, 400 mcg in centres A and B, 50, 50, 200, 300, 600 mcg in centre C and 36.3, 72.6, 108.9, 290.4 and 508.2 mcg in centre D. Challenge was stopped when either the FEV_1_ measured after each inhalation of methacholine decreased more than 20% from baseline or the final dose was given. A test was considered positive if a ≥20% decline in FEV1 from baseline was observed at any dose in centres A, B, and D, or if the total cumulative dose to achieve the decline was ≤1000 mcg in centre C.

*Allergy tests*

Allergy tests were conducted either by skin prick test or specific IgE measurement by ImmunoCAP® to selected allergens or by ImmunoCAP® Rapid (Wheeze/Rhinitis Child set, including house dust mite, wall pellitory, olive pollen, dog dander, timothy, mugwort, birch, cat dander, egg while and cow’s milk). Skin-prick test was considered positive if the allergen wheal size was ≥3 mm, the positive control (histamine) wheal size was ≥3 mm and the negative control (saline) wheal size was <3 mm.

Supplementary table 1. Main reasons for deviations from the standardised diagnostic pathways, categorised by stage of deviation

| **Stage of deviation** | **Reason** | **Practice related** | **Patient related** | **Logistical** | **Unclear** |
| --- | --- | --- | --- | --- | --- |
| No further test after Step 1, n = 14 | Challenge test not due to clinic protocol restriction of age | 4 |  |  |  |
|  | Challenge test not done due to current infection |  | 1 |  |  |
|  | Poor spirometry technique or limited patient cooperation |  | 8 |  |  |
|  | Challenge test not done due to time constraints |  |  | 1 |  |
| Step2b: Bronchial challenge test (or medication trial) taken instead of Step 2a BDR in Pathway 1, n=13 | Poor spirometry technique or limited patient cooperation |  | 3 |  |  |
|  | BDR not done due to supranormal baseline spirometry | 6 |  |  |  |
|  | BDR not done due to time constraints |  |  | 1 |  |
|  | BDR not done since patient was referred with request to conduct challenge test | 1 |  |  |  |
|  | Unclear from medical records |  |  |  | 2 |
| Bronchial challenge test done after a positive BDR in Pathway 1, n=2 | BDR was not considered positive with possible learning effect | 2 |  |  |  |
| No further tests after negative BDR and suspicion of asthma, n=13 | BDR considered positive by change in flow volume curve shape or in body plethysmography parameters | 2 |  |  |  |
|  | Challenge test not done due to appointment availability |  |  | 10 |  |
|  | Challenge test not due to clinic protocol restriction of age | 1 |  |  |  |
| Bronchial challenge test done after abnormal spirometry in Pathway 2, n = 5 | Exercise challenge test done due to exercise related symptoms | 4 |  |  |  |
|  | Spirometry considered not obstructive with consideration to FVC and FEV_1_/FVC | 1 |  |  |  |
| BDR done after normal spirometry in Pathway 2, n =10 | BDR considered more feasible than MCT due to poor spirometry technique |  | 5 |  |  |
|  | Spirometry considered obstructive by MMEF_25-75_ value | 1 |  |  |  |
|  | Flow volume curve considered concave at the time of spirometry and BDR done, but later interpreted as normal by clinician review | 2 |  |  |  |
|  | Challenge test not done due to current infection |  | 1 |  |  |
|  | Unclear from medical record |  |  |  | 1 |
| **Total (n=57)** |  | **24** | **18** | **12** | **3** |

BDR: bronchodilator reversibility, MCT: methacholine challenge test
